# Longitudinal experiences and impact of the COVID-19 pandemic among people with past or current eating disorders in Sweden

**DOI:** 10.1101/2021.03.29.21254526

**Authors:** Andreas Birgegård, Afrouz Abbaspour, Stina Borg, David Clinton, Emma Forsén Mantilla, Jet D. Termorshuizen, Cynthia M. Bulik

**Affiliations:** Department of Medical Epidemiology and Biostatistics, Karolinska Institutet, Stockholm, Sweden; Rivierduinen Eating Disorders Ursula, Leiden, The Netherlands; Department of Psychiatry, University of North Carolina at Chapel Hill, Chapel Hill, North Carolina, USA; Department of Nutrition, University of North Carolina at Chapel Hill, Chapel Hill, North Carolina, USA

**Keywords:** eating disorders, pandemic, COVID-19, social isolation, relapse

## Abstract

**Objective:** To document the impact of the COVI-19 pandemic on the health and well-being of individuals with past and current eating disorders in Sweden.

**Method:** We re-contacted participants from two previous Swedish studies who had a known lifetime history of an eating disorder. Participants completed an online questionnaire about their health and functioning at baseline early in the pandemic (Wave 1; *N*=982) and six months later (Wave 2); *N*=646).

**Results:** Three important patterns emerged: 1) higher current eating disorder symptom levels were associated with greater anxiety, worry, and pandemic-related eating disorder symptom increase; 2) patterns were fairly stable across time, although a concerning number who reported being symptom-free at Wave 1 reported re-emergence of symptoms at Wave 2; and only a minority of participants with current eating disorders were in treatment, and of those who were in treatment, many reported fewer treatment sessions than pre-pandemic and decreased quality of care.

**Conclusions:** The COVID-19 pandemic is posing serious health challenges for individuals with eating disorders, whether currently symptomatic or in remission. We encourage health service providers and patient advocates to be alert to the needs of individuals with eating disorders and to take active measures to ensure access to appropriate evidence-based care both during and following the pandemic.

**Significant Outcomes and Limitations:**

- Individuals with eating disorders symptoms or current active disorder report higher adverse impact of COVID-19 on their mental health
- Even individuals who were symptom-free early in the pandemic reported a resurgence of eating disorder symptoms
- A large proportion of symptomatic individuals were not in treatment for their eating disorder, services should be aware and access to evidence-based care should be ensured across Sweden
- Limitations included the use of a convenience sample with atypical diagnostic distribution, and a low initial response rate, possibly introducing bias and limiting generalisability.

**Data Availability Statement:** Fully anonymized data are available from the corresponding author upon request.

## Introduction

Coronavirus disease 2019 (COVID-19), an infectious disease caused by severe acute respiratory syndrome coronavirus 2 (SARS-CoV-2), was declared a pandemic in March 2020 by the World Health Organization. In response, countries across the globe implemented varyingly strict measures to limit the spread of the virus, balancing the impact of public health measures on social isolation, disruption of daily routines, and economic factors. Besides the direct and often prolonged impact of COVID-19 on physical and mental health,^1,2^ pandemic- related restrictions have also adversely affected mental health in the general population,^34^ and may worsen the symptoms in individuals with pre-existing psychiatric illnesses.^56^ We surveyed a large sample of individuals with eating disorders (ED) in Sweden at two timepoints—Wave 1 (baseline early in the pandemic) and Wave 2 (six months later)—to document the impact of the pandemic and public health measures taken to limit its transmission on individuals with EDs.

Several studies emerged in the literature early suggesting that the pandemic is adversely impacting individuals with ED. Reports from around the world suggest that exposure to triggering environments (e.g., having to spend more time in close quarters with family or roommates), lack of social support, and the absence of structure to daily life are particularly challenging for individuals with EDs.^7,8^ Individuals surveyed report increases in ED symptoms, anxiety, and stress as a result of the pandemic.^9,10^ In addition, disruption to clinical services in many countries has impeded access to treatment and initially, virtual care was rated as less satisfactory than typical face-to-face treatment.^10,11^ Evidence of exacerbation of ED symptoms in individuals with an ED during the COVID-19 pandemic has been documented in studies from, for example, Spain^12^, Australia^13^, Germany^14^, the United States, and the Netherlands.^10^

Our primary aim was to characterize experiences of individuals with a current or past ED in Sweden during the COVID-19 pandemic using a longitudinal design with assessments at baseline (Wave 1) and after 6 months (Wave 2). Sweden-specific information is valuable as the country has had a unique response to COVID-19 public health management. We investigated changes in illness status over time, ED symptoms, anxiety, treatment availability, and COVID- 19-related concerns. We also investigated baseline variables that were associated with deterioration or improvement of illness status over time. Our results provide valuable information for patients and families, clinicians, and advocacy groups about needs of individuals with EDs in Sweden and have the potential to guide care strategies and resource planning during this and future pandemics.

## Material and Methods

### Participants and procedure

We contacted all participants from two previous large-scale ED studies in Sweden: the Anorexia Nervosa Genetics Initiative (ANGI)^15^ and the Binge Eating Genetics Initiative (BEGIN), who had given permission to be recontacted for future research. All participants had lifetime history of an ED (and many had current ED). Although all ED presentations were included, the majority of participants had anorexia nervosa (AN), given the nature of the parent ANGI study. Although all participants had verified diagnoses from the parent studies, for the purposes of this study, we used self-reported diagnosis and self-reported symptom status. We sent 3,774 emails on May 27^th^ (170 emails were undeliverable) and when we froze first-wave data collection after six weeks, 982 individuals had responded (27%). The second wave, sent only to those who had completed the Wave 1 survey, was administered between December 22^nd^ 2020 and February 2^nd^ 2021, and 646 responded (66% of Wave 1). The study methodology was approved by the Swedish Ethical Review Authority (Dnr 2020-04136).

### Survey

The survey (see Supplement 1) was modelled after a survey used in the United States and the Netherlands to study the impact of the pandemic on individuals with EDs (Termorshuizen et al.^10^) that queried physical and mental well-being related to COVID-19 in the previous two weeks. Participants provided data on age, sex, gender identity, diagnostic and treatment status, exposure to COVID-19, and current situational circumstances (e.g., quarantined, physical distancing). A Likert scale was used to measure level of concern about changes in ED symptoms, frequency of symptoms, and worry related to COVID-19. The 7-item version of the Generalised Anxiety Disorder Scale (GAD-7)^16^ assessed anxiety, with Cronbach’s alpha of .91 at Wave 1 and .92 at Wave 2. We applied the ≥10 cutoff^16^ for possible diagnosable generalised anxiety disorder to categorize participants. Free-text items invited participants to share additional comments (qualitative analyses will be discussed elsewhere).

### Statistical analysis

We defined three subgroups according to self-reported symptom status, with the question “Which of the following statements best describes your experience?” and response options “I have previously had an ED but am currently free of symptoms” (**NoSx**), “I have previously had an ED and still experience some symptoms” (**Sx**), and “I currently have an ED” (**ED**). Statistics for each wave are based on the symptom grouping reported at that timepoint, whereas longitudinal analyses (migration) are based on Wave 1 groupings. Distribution at both time points and migration across symptom groups over time are shown in Figure 1.

**Figure 1.**
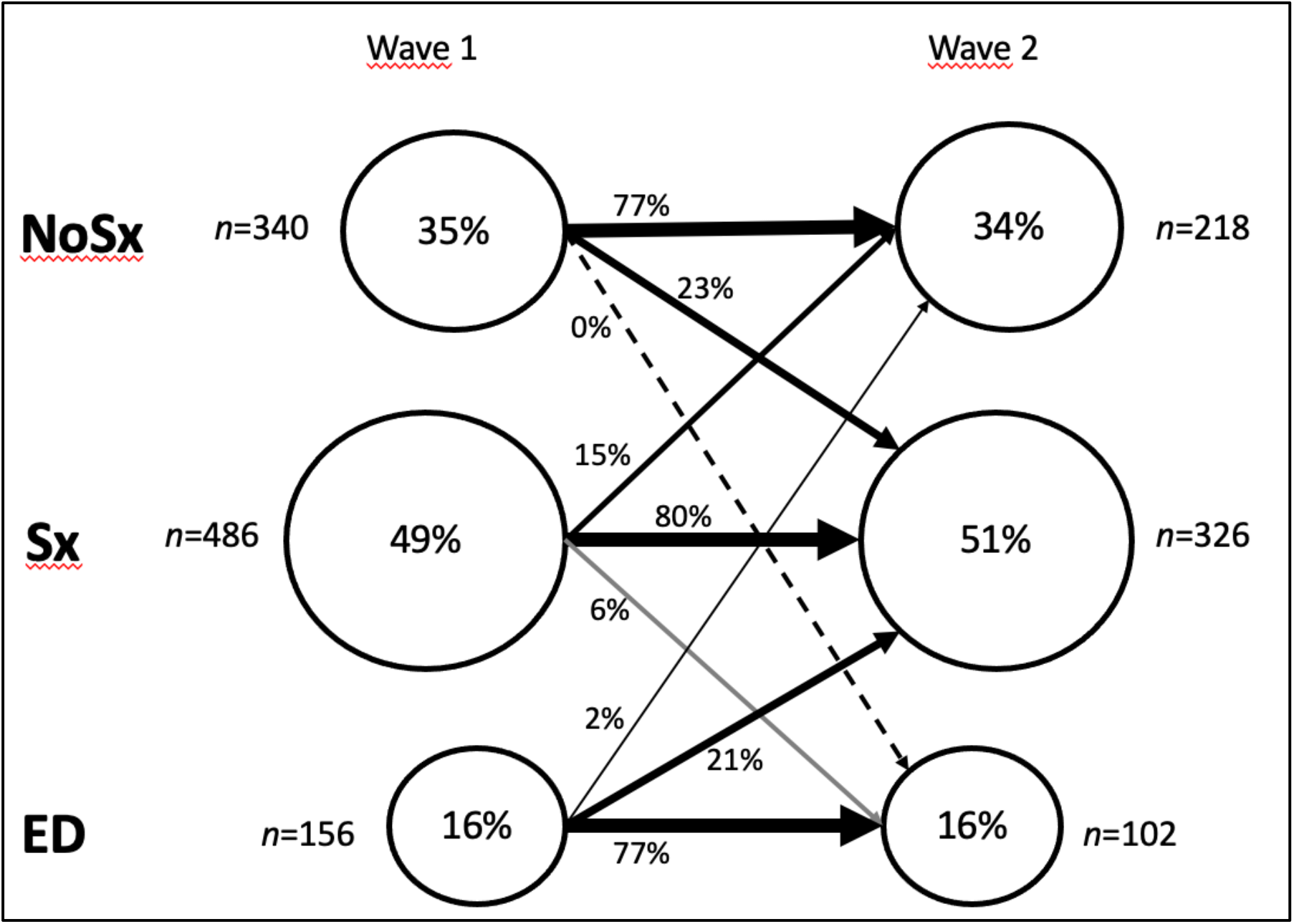
Distribution (*n*, %) of participants into self-reported symptom-level groups at each measurement wave, where **NoSx**=previous ED but no current symptoms, **Sx**=previous ED and remaining symptoms, and **ED**=current ED. Circle sizes approximate group size, and migration between groups (arrows) is displayed with percentage of starting group moving to another.

**Figure 2.**
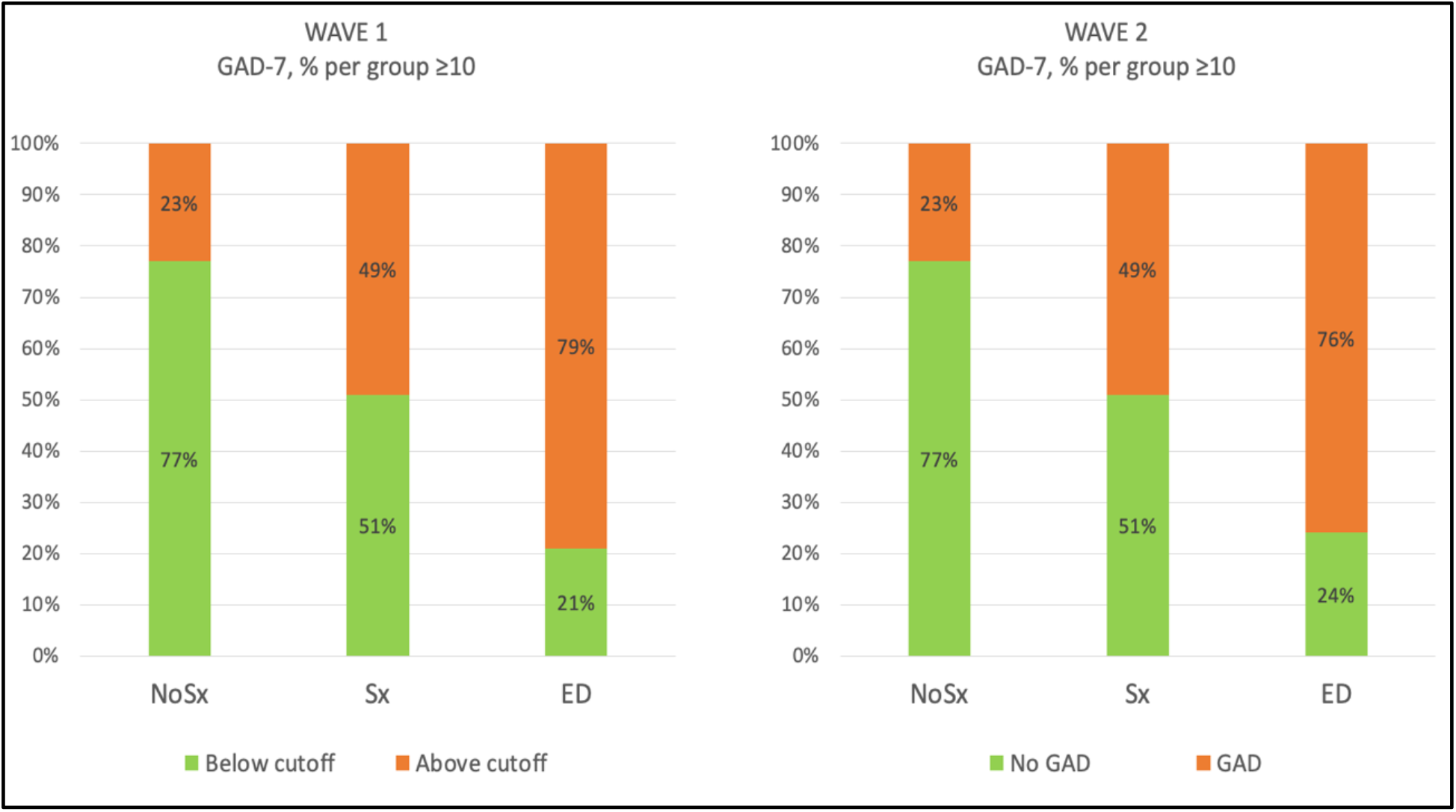
Proportion of individuals scoring above/below the GAD-7 cut-off (≥10) at each time wave, separated by ED status group.

We examined changes in each symptom status group over time and baseline predictors of symptom deterioration or improvement using Welch’s *t*-tests (due to imbalanced design) and Cohen’s *d* effect sizes, by comparing those who changed group over time with baseline group peers who did not change. Three contrasts were possible based on statistical power: symptom deterioration (**NoSx**→**NoSx** vs. **NoSx**→**Sx**), and two comparisons marked by symptom improvement (**Sx**→**Sx** vs. **Sx**→**NoSx** and **ED**→**ED** vs. **ED**→**Sx**). We selected baseline predictors that captured ED symptoms, anxiety level (GAD-7), and items that captured concerns about factors that might increase ED symptoms.

Attrition analyses compared Wave 2 responders with those who responded only at Wave 1 to investigate representativeness of the longitudinal subsample, on anxiety, worry about symptom increase, and ED symptoms.

## Results

### Sample characteristics and distribution of ED symptom groups

The sample consisted of 98% biological females, and gender identity distribution was 97% female, 2% male, and 1% non-binary or other. Self-reported previous or current ED diagnosis distribution (participants could mark several response alternatives) was: anorexia nervosa 64%; bulimia nervosa 37%; binge-eating disorder 24%; and other specified feeding and eating disorder 45%; with 12% other; and 1% responding “Don’t know/prefer not to answer”. Table 1 shows descriptive statistics for COVID-19-related exposure and preventive measures at Wave 1 and Wave 2. No significant differences emerged between individuals who responded to both waves of data collection and those who responded to Wave 1 only on any tested baseline variables.

**Table 1.** COVID-19 exposure and preventive measures; % “Yes” responses (See Supplement for full questionnaire)

| Item | Wave 1 *n* (%) | Wave 2 *n* (%) |
| --- | --- | --- |
| Practicing social distancing | 749 (77%) | 556 (87%) |
| Forced isolation | 10 (1%) | 3 (1%) |
| Work/study from home | 457 (47%) | 341 (53%) |
| Ordered home stay | 51 (5%) | 37 (6%) |
| Tested positive for COVID-19 | 17 (2%) | 56 (9%) |
| Clinical COVID-19 diagnosis | 6 (1%) | 4 (1%) |
| Have not had COVID-19 | 618 (63%) | 443 (69%) |
| Maybe had COVID-19 but not sure | 336 (34%) | 139 (22%) |

Figure 1 shows the distribution of the symptom groups **NoSx, Sx**, and **ED** at each time point. Thirty-four percent had no ED symptoms, ∼50% had lingering symptoms, and 16% reported a current active ED at each time point. Although the percentages at each timepoint were very similar, migration did occur. A full 23% of individuals transitioned from **NoSx** at Wave 1 to **Sx** at Wave 2 (i.e., deterioration); 15% of individuals with **Sx** at baseline reported **NoSx** at Wave 2 (i.e., improvement); and 21% of individuals with **ED** at Wave 1 reported **Sx** at Wave 2 (i.e., improvement). The majority of individuals, however, remained in the same symptom category across time.

### Anxiety and ED symptom variables by symptom status group

Table 2 reveals that individuals in all three symptoms groups were more likely to say that their anxiety had increased since the end of 2019 at Wave 2 than at Wave 1. In most cases across symptom groups and at both waves, participants were more likely to attribute their increases in anxiety to COVID-19 (data not shown).

**Table 2.**
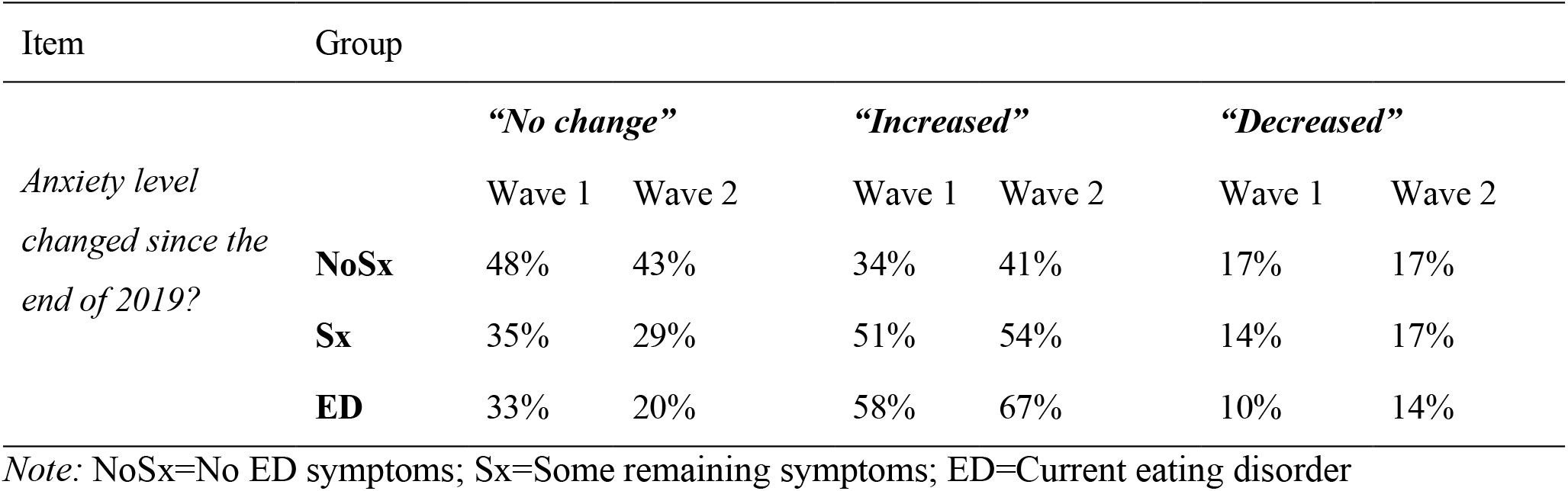
Changes in self-reported anxiety by symptom group.

| Item | Group | *“No change”* |  | *“Increased”* |  | *“Decreased”* |  |
| --- | --- | --- | --- | --- | --- | --- | --- |
|  |  | Wave 1 | Wave 2 | Wave 1 | Wave 2 | Wave 1 | Wave 2 |
| *Anxiety level changed since the end of 2019?* | **NoSx** | 48% | 43% | 34% | 41% | 17% | 17% |
|  | **Sx** | 35% | 29% | 51% | 54% | 14% | 17% |
|  | **ED** | 33% | 20% | 58% | 67% | 10% | 14% |

*Note:* NoSx=No ED symptoms; Sx=Some remaining symptoms; ED=Current eating disorder

GAD-7 was strongly associated with symptom group, and patterns were similar across waves. At both Wave 1 and Wave 2, 23% of the **NoSx** group, ∼50% of the **Sx** group, and 79% (Wave 1) and 76% (Wave 2) of the **ED** group scored above the cut-off for GAD-7.

Table 3 shows that concern about ED symptom increase due to the pandemic^1^ was strongly related to symptom status group. Participants in **ED** were more concerned about increased symptoms due to lack of structure and social support, and being exposed to triggering environments, than **Sx**, who in turn were more concerned than **NoSx**. Patterns were similar across waves.

**Table 3.** Worry about impact of COVID-19 by symptom group and measurement wave for items related to concern about factors leading to increased ED symptoms, ED symptoms themselves, and worry about the impact of COVID-19 on others and own health. See Supplement for items.

| Variable | Response options shown | Symptom group | Wave 1 | Wave 2 |
| --- | --- | --- | --- | --- |
| Worry that ED symptoms increase due to lack of structure | Per cent responding<br>“Fairly worried” to<br>“Very worried” | NoSx | 10% | 7% |
|  |  | Sx | 33% | 31% |
|  |  | ED | 56% | 61% |
| Worry that ED symptoms increase due to lack of social support |  | NoSx | 3% | 4% |
|  |  | Sx | 20% | 19% |
|  |  | ED | 51% | 48% |
| Worry that ED symptoms increase due to being in a triggering environment |  | NoSx | 9% | 6% |
|  |  | Sx | 32% | 32% |
|  |  | ED | 56% | 53% |
| Binge eating bunkered food | Per cent responding<br>“Often” to “Daily or more” | NoSx | 1% | 1% |
|  |  | Sx | 8% | 7% |
|  |  | ED | 24% | 23% |
| Restricted food intake due to COVID-19-related factors |  | NoSx | 5% | 2% |
|  |  | Sx | 14% | 11% |
|  |  | ED | 27% | 24% |
| Compensated for food intake food intake due to COVID-19-related factors |  | NoSx | 1% | 1% |
|  |  | Sx | 9% | 8% |
|  |  | ED | 26% | 28% |
| Worried about not being able to exercise |  | NoSx | 17% | 18% |
|  |  | Sx | 36% | 37% |
|  |  | ED | 39% | 46% |
| Worry about self being infected | Per cent responding<br>“Fairly worried” to<br>“Very worried” | NoSx | 23% | 39% |
|  |  | Sx | 28% | 38% |
|  |  | ED | 32% | 40% |
| Worry about others being infected |  | NoSx | 79% | 79% |
|  |  | Sx | 78% | 84% |
|  |  | ED | 80% | 86% |
| Worry about physical health being affected |  | NoSx | 28% | 39% |
|  |  | Sx | 37% | 50% |
|  |  | ED | 45% | 54% |
| Worry about mental health being affected |  | NoSx | 42% | 50% |
|  |  | Sx | 59% | 65% |
|  |  | ED | 65% | 73% |

*Note:* NoSx=No ED symptoms; Sx=Some remaining symptoms; ED=Current eating disorder

Table 3 further shows that more participants reported being worried about others being infected by COVID-19 than themselves, and increased ED symptom level status was associated with higher worry. Further, participants were much more worried about their mental health being affected than physical health, and there was a clear pattern of higher ED symptoms being associated with higher levels of concern.

A similar pattern emerged with ED symptoms, with increasing symptoms of binge eating, restrictive eating, and compensatory behaviours across **NoSx, Sx**, and **ED** groups. Concern about not being able to exercise was similar in both **ED** and **Sx**. Patterns were similar across waves.

### Treatment

Table 4 presents treatment-related results. Notably, the majority of individuals with active EDs reported not being in current treatment. Most notably, a majority had no current ED treatment, 40% had had fewer sessions in the last two weeks, and a substantial proportion experienced their treatment quality as worse or much worse than before the pandemic. On the other hand, fairly many still had face-to-face sessions and a majority experienced their treatment as good or better than before the pandemic.

**Table 4.** Experiences of treatment.

| Item | “F2F sessions” |  | “Transitioned to virtual” |  | “No sessions” |  | “Not in treatment” |  |
| --- | --- | --- | --- | --- | --- | --- | --- | --- |
| Presence and format of treatment | Wave 1 | Wave 2 | Wave 1 | Wave 2 | Wave 1 | Wave 2 | Wave 1 | Wave 2 |
|  | 27% | 28% | 10% | 9% | 7% | 13% | 58% | 51% |
| Change in session frequency | “Fewer sessions” |  | “As many sessions” |  | “More sessions” |  |  |  |
|  | Wave 1 | Wave 2 | Wave 1 | Wave 2 | Wave 1 | Wave 2 |  |  |
|  | 40% | 41% | 53% | 54% | 7% | 5% |  |  |
| Change in ED treatment quality | “Better” |  | “As good” |  | “A little worse” |  | “Much worse” |  |
|  | Wave 1 | Wave 2 | Wave 1 | Wave 2 | Wave 1 | Wave 2 | Wave 1 | Wave 2 |
|  | 4% | 5% | 56% | 49% | 29% | 30% | 11% | 16% |

*Note:* F2F=Face to face; ED=eating disorder. Responses regarding treatment were reported for the ED group only (Wave 1 *n*=156, Wave 2 *n*=102). Responses regarding change in session frequency and treatment quality were asked only to those who responded to have had face-to-face or virtual sessions in the past two weeks (Wave 1 *n*=55, Wave 2 *n*=37).

### Migration between symptom status groups

We conducted two analyses exploring Wave 1 predictors of a change in symptom status between Wave 1 and Wave 2 (Table 5). Two analyses investigated factors associated with deterioration and two related to improvement.

**Table 5.** Descriptive statistics, Welch’s t-tests and Cohen’s d effect sizes of the three migration analyses using baseline variables; Stable **NoSx** vs. **NoSx**→**Sx** (n=168 vs. 51), Stable **Sx** vs. **Sx**→**NoSx** (n=252 vs. 45), and Stable **ED** vs. **ED**→**Sx** (n=84 vs. 23). See Supplementary 1 for item wording.

| Compared groups | GAD-7 | Worry about mental health being affected | Concern about lack of structure | Concern about lack of social support | Concern about triggering environment | Binge eating | Restrictive eating | Compensatory symptoms (e.g. vomiting) | Worry about no exercise |
| --- | --- | --- | --- | --- | --- | --- | --- | --- | --- |
|  | M (SD) | M (SD) | M (SD) | M (SD) | M (SD) | M (SD) | M (SD) | M (SD) | M (SD) |
| *Stable NoSx* | 6.0 (4.24) | 1.7 (.78) | 1.4 (.68) | 1.2 (.42) | 1.3 (.58) | 1.1 (.27) | 1.2 (.52) | 1.1 (.30) | 1.6 (.83) |
| vs. *NoSx→Sx* | ***, †† |  | *; † | *; † | ** ; †† |  |  |  | ** ; †† |
|  | 9.2 (5.60) | 1.8 (.67) | 1.7 (.88) | 1.4 (.64) | 1.8 (1.02) | 1.2 (.43) | 1.3 (.68) | 1.1 (.35) | 2.1 (1.04) |
| *Stable Sx* | 10.2 (5.43) | 1.8 (.68) | 2.2 (.97) | 1.8 (.96) | 2.1 (1.02) | 1.4 (.64) | 1.5 (.76) | 1.4 (.65) | 2.1 (1.09) |
| vs. *Sx→NoSx* |  |  | ** ; † | ** ; † | * ; † |  |  | * ; † |  |
|  | 9.4 (5.62) | 1.8 (.64) | 1.8 (.84) | 1.4 (.69) | 1.7 (.92) | 1.3 (.64) | 1.5 (.76) | 1.2 (.46) | 2.0 (1.02) |
| *Stable ED* | 14.5 (4.95) | 1.8 (.58) | 2.7 (1.13) | 2.6 (1.13) | 2.6 (1.07) | 1.6 (.94) | 1.7 (.94) | 1.7 (.96) | 2.1 (1.11) |
| vs. *ED→Sx* | * ; †† |  |  |  |  |  |  |  |  |
|  | 11.6 (4.98) | 1.6 (.66) | 2.6 (1.23) | 2.4 (1.11) | 3.0 (1.07) | 2.0 (1.04) | 1.5 (.73) | 1.7 (.94) | 2.3 (1.02) |

\* $p < .05$ ; \*\* $p < .01$ ; \*\*\* $p < .001$ . † $d \geq .20$ (small effect); †† $d \geq .50$ (medium effect).
*Note:* NoSx=No ED symptoms; Sx=Some remaining symptoms; ED=Current eating disorder; GAD-7=Generalised anxiety disorder 7-item scale; ICB=name of the ED symptom items.

Those individuals who transitioned from **NoSx** to **Sx** had significantly higher Wave 1 mean GAD-7 scores, greater concern about being in a triggering environment, and higher worry about not being able to exercise, with medium effect sizes. They also had significantly higher concern about lack of structure and social support, but with small effect sizes.

Individuals who transitioned from **Sx** at Wave 1 to **No Sx** at Wave 2 had lower scores on concerns about lack of structure, social support, and being in a triggering environment and lower scores on compensatory behaviours, with small to medium effects. Individuals who moved from **ED** at Wave 1 to **Sx** at Wave 2 had lower baseline GAD-7 scores.

## Discussion

We characterized experiences of people with a current or past ED early in the COVID-19 pandemic and six months later. Using a survey coordinated with studies in the USA and the Netherlands, we identified three important patterns. First, the higher the current ED symptom level of the participant, the more anxiety, worry, and ED symptom increase was reported. Second, results were fairly stable across time, with some exceptions. Third, quite concerningly, only a minority of participants with current ED were in treatment, and of those who were in treatment many reported fewer treatment sessions than usual and decreased quality of care.

The overall impression is that among people with experience of an ED, those with lingering symptoms or current active disorder are particularly vulnerable to the disruptions and anxiety caused by the COVID-19 pandemic and the societal restrictions that have necessarily been put in place. GAD-7 results suggested that a full three-quarters of individuals with active EDs were likely to also have generalised anxiety disorder. In the absence of pre-pandemic data, we are unable to determine whether this is higher than would be expected, but regardless it reflects the importance of attending to anxiety in the treatment of individuals with EDs during the pandemic. Although we found relatively few relapses into ED, a concerning number who initially reported being symptom-free reported re-emergence of symptoms as the pandemic progressed.

Regarding predictors of deteriorating symptoms, baseline anxiety, weaker social support and structure, and fears about not being able to exercise were all associated with worsening ED symptoms. Symptom improvement showed to some extent the opposite pattern, as it was associated with lower anxiety and higher ratings on social support and structure. These results may be especially informative for treatment planning and safeguarding continued recovery, since they underscore the importance of ensuring that individuals with current or past EDs have skills to address anxiety and strategies to forge social connectedness as well as support and adequate structure to their daily lives.

Intriguingly, our results closely mirror results reported in the US and the Netherlands,^10^ suggesting that despite considerable differences in the public health measures used across the three countries to contain transmission of COVID-19, the impact on individuals with EDs was comparable. Indeed, EDs thrive in social isolation with patients reporting feeling alone with their ED thoughts and isolated in terms of monitoring their weight and eating. Since developing regular and adequate eating is central to ED recovery and maintenance of treatment gains, the absence of a predictable daily structure removes a central component of ED care.

Our results have direct clinical implications for clinicians working with ED patients during the pandemic as well as for advocacy organizations who provide additional support. Even if patients have been transitioned to virtual care, it is important to ensure that they have adequate social support via families, peers, advocacy organizations, or even online forums to maintain accountability and motivation for recovery. It is also essential to assess directly individuals’ living arrangements. Given limitations on socializing and recommendations for physical distancing during the pandemic, individuals may find themselves with less freedom and flexibility to extricate themselves from unhelpful or even toxic environments. Remaining vigilant for triggering situations and providing practical assistance with managing them is of critical import. Finally, assisting patients with developing and maintaining structure to their daily lives, especially when working from home, can aid recovery. Providing support, social connectedness, and daily structure are three clinical targets that appear to be critical to ED patients during the pandemic.

Although our study provides important knowledge about how individuals with ED are coping with the pandemic, it has several limitations. First, given our intention to field a survey soon after COVID-19 was declared a global pandemic, we rapidly translated and adapted a survey used in the US and the Netherlands. This precluded typical steps to ensure sound psychometric properties. Second, we relied on a convenience sample (i.e., individuals who had participated in previous studies who agreed to be contacted for future research), which did not reflect the distribution of diagnoses to be expected from a community sample and may have biased results. Third, we relied on self-report diagnosis and symptom reports to characterize patients. Although all participants had formal diagnoses in the past, the use of self-reports to establish current status remains a limitation. Fourth, our response rate to the initial survey was low (27%) and we experienced attrition between the waves, again potentially introducing bias and limiting generalisability. Finally, we were underpowered for some analyses; group sizes were low for analysing transitions between symptom levels across time.

Nevertheless, we were able to capture fairly early on in the pandemic how individuals with ED were being affected. As anticipated, COVID-19 has been particularly challenging for those with existing active mental disorders, and even those who are currently symptom-free are challenged to remain healthy. We encourage clinicians and advocacy organizations to avail themselves of these results to aid in developing resources for patients, families, and clinicians for dealing with ED during the pandemic. Given the frequency with which individuals with active EDs reported not being in treatment and the concerning number who initially reported being symptom-free who subsequently reported re-emergence of symptoms six months later, we encourage health service providers and patient advocates to be alert to the needs of ED patients and to take active measures to ensure access to appropriate evidence-based care for all EDs across Sweden during the pandemic and subsequently when there may be a serious backlog of individuals requiring care.

## Supporting information

Survey

## Data Availability

Fully anonymized data are available from the corresponding author upon request.

## Acknowledgements

Dr Bulik acknowledges support from the Swedish Research Council (Vetenskapsrådet, award: 538-2013-8864). The authors also thank Peter Lind for valuable support with IT.

## Footnotes

1 Some response options in this item (“concern”, see Supplement 1) are not shown since scores were very low as the response options are relatively irrelevant for Swedish conditions; these related to concern about not being able to afford food or treatment, which is not an issue in Sweden at this point.

