## Supplementary material for "Longitudinal experiences and impact of the COVID-19 pandemic among people with past or current eating disorders in Sweden": Survey

**ed_dx**
Which of the following eating disorders do you currently have or have you had in the past? (Please mark all that apply)

- ❑ Anorexia nervosa (1)
- ❑ Bulimia nervosa (2)
- ❑ Binge-eating disorder (3)
- ❑ Avoidant restrictive food intake disorder (ARFID) (4)
- ❑ Atypical anorexia nervosa (5)
- ❑ Purging disorder (6)
- ❑ Night-eating syndrome (7)
- ❑ Other specified feeding or eating disorder (OSFED or EDNOS) (8)
- ❑ Other eating disorder (9)
- ❑ Don't know/prefer not to answer (10)
- ❑ I have never had an eating disorder (11)

**ed_exp**
Which of the following best describes your experience?

- ❍ I had an eating disorder in the past and have no current symptoms. (1)
- ❍ I had an eating disorder in the past and still have some lingering symptoms. (2)
- ❍ I currently have an eating disorder. (3)

**gender**
What is your gender identity?

- ❍ Man (1)
- ❍ Woman (2)
- ❍ Nonbinary / gender fluid (3)
- ❍ Other (4)

**sex**
What was your biological sex at birth?

- ❍ Male (1)
- ❍ Female (2)
- ❍ Intersex (3)

**ort**
What is your area code?
*Please answer 5 numbers without space*

**anx2019**
Do you think your anxiety levels have changed since the end of 2019?

- ❍ No (1)
- ❍ Yes - my levels of anxiety have increased (2)
- ❍ Yes - my levels of anxiety have decreased (3)

**anx_covid**
How much do you think these changes are due to the COVID-19 (coronavirus) situation?
- ❍ Not at all (1)
- ❍ Somewhat (2)
- ❍ A lot (3)

**distance**
The following question relates to the COVID-19 (coronavirus) situation. Are you currently...?

| | Yes (1) | No (2) |
|---|---|---|
| Quarantined (1) | ○ | ○ |
| Practicing physical distancing (also known as social distancing) (2) | ○ | ○ |
| In voluntary self-isolation (3) | ○ | ○ |
| In mandatory self-isolation (4) | ○ | ○ |
| Working from home (5) | ○ | ○ |
| Shelter-in-place/stay-at-home order (6) | ○ | ○ |

**covid_exp**
During the last **two weeks**, have you been exposed to someone likely to have coronavirus (COVID-19)?
- ❍ Yes, with positive COVID-19 test (1)
- ❍ Yes, with medical diagnosis (2)
- ❍ Yes, but not diagnosed (3)
- ❍ No (4)
- ❍ Unknown/unsure (5)

**covid_dx**
Have **you** been diagnosed with coronavirus (COVID-19)?
- ❍ Yes, positive COVID-19 test (1)
- ❍ Yes, medical diagnosis (2)
- ❍ No (3)
- ❍ Possibly, but I was not diagnosed or tested (4)

**covid_sxs**
Have you had any of the following symptoms in the past month? Please select all that apply.
- ❑ Fever (1)
- ❑ Cough (2)
- ❑ Shortness of breath (3)
- ❑ Sore throat (4)
- ❑ Fatigue (5)
- ❑ Loss of taste or smell (6)

**covid_fam**
Has anyone in your family been diagnosed with coronavirus (COVID-19)?
- ❍ Yes, member of household (1)
- ❍ Yes, a family member who does not live with me (2)
- ❍ No (3)

**covid_event**

Have any of the following happened to your family members because of coronavirus (COVID-19)?

- ❑ Become ill physically (1)
- ❑ Hospitalized (2)
- ❑ Isolated/put into quarantine (3)
- ❑ Lost job (4)
- ❑ Other (5)
- ❑ No, none of the above (99)

**covid_event_oth**

Please describe what else happened to your family due to the coronavirus (COVID-19):

**covid_worry**

How worried are you...

| | Not worried at all (1) | (2) | (3) | Somewhat worried (4) | (5) | (6) | Very worried (7) |
|---|---|---|---|---|---|---|---|
| ...about being infected **yourself?** (1) | ○ | ○ | ○ | ○ | ○ | ○ | ○ |
| ...about **others** being infected? (2) | ○ | ○ | ○ | ○ | ○ | ○ | ○ |
| ...that your **physical health** could be influenced by coronavirus (COVID-19)? (3) | ○ | ○ | ○ | ○ | ○ | ○ | ○ |
| ...that your **mental health** could be influenced by coronavirus (COVID-19)? (4) | ○ | ○ | ○ | ○ | ○ | ○ | ○ |

**covid_thought**

| | Not at all (1) | (2) | (3) | About half the day (4) | (5) | (6) | All day long (7) |
|---|---|---|---|---|---|---|---|
| How much of your day are you thinking about COVID-19? (1) | ○ | ○ | ○ | ○ | ○ | ○ | ○ |

**covid_pos**

| | Not at all (1) | (2) | (3) | Some positive changes (4) | (5) | (6) | Several positive changes (7) |
|---|---|---|---|---|---|---|---|
| Has the COVID-19 situation led to any positive changes in your life? (1) | o | o | o | o | o | o | o |

**covid_pos2**
Please describe any positive changes.

________________________________________________________________

**concern**
Please answer the following questions based on the past **two weeks.**
I have been concerned about...

| | Not at all concerned (1) | Slightly concerned (2) | Somewhat concerned (3) | Very concerned (4) |
|---|---|---|---|---|
| ... having access to enough food (e.g., unable to go to a grocery store regularly, unable to leave home, etc.). (1) | o | o | o | o |
| ... accessing foods that are consistent with my current meal plan/style of eating. (2) | o | o | o | o |
| ... worsening of my eating disorder due to a lack of structure. (3) | o | o | o | o |
| ... worsening of my eating disorder due to a lack of social support. (4) | o | o | o | o |
| ... worsening of my eating disorder due to increased time living in a triggering environment. (5) | o | o | o | o |
| … being able to afford the food I need for recovery due to loss of income related to COVID-19. (6) | o | o | o | o |
| … being able to afford eating disorder treatment due to loss of income related to COVID-19. (7) | o | o | o | o |

**ICB**

Please answer the following questions based on the past **two weeks**.
In the past **two weeks**, I have...

| | Not at all (1) | Once or twice (2) | Frequently (3) | Daily or more (4) |
|---|---|---|---|---|
| … binged on food that I (or my family or roommate) have stockpiled. (1) | ○ | ○ | ○ | ○ |
| … restricted my intake more because of COVID-19-related factors. (2) | ○ | ○ | ○ | ○ |
| … engaged in more compensatory behaviors (e.g., self-induced vomiting, excessive exercise, misuse of laxatives and/or water pills) because of COVID-19-related factors. (3) | ○ | ○ | ○ | ○ |
| ...felt anxious about not being able to exercise. (4) | ○ | ○ | ○ | ○ |

**oth_concern**

In the last **two weeks**, what other eating disorder-related concerns have you had that are not listed above?

________________________________________________________________

**good_change**

In the past two weeks, have you experienced any positive changes in your eating disorder symptoms?

❍ Yes (1)
❍ No (2)

**good_change_txt**

Please describe those changes:

________________________________________________________________

**treatment**

Choose the alternative that best characterizes your situation during the **past two weeks**.

In the last **two weeks**:

❍ I have had face-to-face (in person) interactions with my eating disorders treatment provider(s). (1)
❍ I have transitioned to online care with my eating disorders treatment provider(s) (i.e., telehealth) (2)
❍ I have not been able to engage with my eating disorders treatment provider(s) at all. (3)
❍ I do not currently receive eating disorders treatment. (4)

**Q85**

In the last **two weeks:**

❍ I have had to reduce the number of sessions/contacts with my eating disorders treatment provider(s). (1)
❍ I have had at least the same number of sessions/contacts with my eating disorders treatment provider(s). (2)

**trt_quality**
The quality of my treatment in the **past two weeks** has been…
- ❍ Better than usual (1)
- ❍ As good as usual (2)
- ❍ Somewhat worse than usual (3)
- ❍ Much worse than usual (4)

**trt_need**
[*Not required*]
In the last **two weeks**, what have been your greatest needs with regard to eating disorder treatment or support?

________________________________________________________________

**gad7**
Over the last 2 weeks, how often have you been bothered by the following problems?

| | Not at all (1) | Several days (2) | Over half the days (3) | Nearly every day (4) |
|---|---|---|---|---|
| Feeling nervous, anxious or on edge (1) | ○ | ○ | ○ | ○ |
| Not being able to stop or control worrying (2) | ○ | ○ | ○ | ○ |
| Worrying too much about different things (3) | ○ | ○ | ○ | ○ |
| Trouble relaxing (4) | ○ | ○ | ○ | ○ |
| Being so restless that it's hard to sit still (5) | ○ | ○ | ○ | ○ |
| Becoming easily annoyed or irritable (6) | ○ | ○ | ○ | ○ |
| Feeling afraid as if something awful might happen (7) | ○ | ○ | ○ | ○ |

**anx_impact**
If you checked off any problems, how difficult have these made it for you to do your work, take care of things at home, or get along with other people?
- ❍ Not difficult at all (1)
- ❍ Somewhat difficult (2)
- ❍ Very difficult (3)
- ❍ Extremely difficult (4)
